# Targeted Proteomic Profiling of Nasal Fluid from the Brain-Nose Interface

**DOI:** 10.64898/2026.06.16.26355519

**Authors:** Mohammad Bashiri, Maria Babu, Gunter Weiss, Stefan Kotschote, Richard Metzler, Hilary Wunderlich, Agnese Petrera, Elena De Domenico, Heidi Theis, Katrin Lehmann, Frederik Heinrich, Mir-Farzin Mashreghi, Marion San Nicoló, Mareike Albert, Sabine Mertzig

**Author notes:** **Corresponding author** Dr. Sabine Mertzig, Noselab GmbH, Widenmayerstrasse 27, 80538, Munich.

## Abstract

The brain-nose interface is an anatomical junction where olfactory neurons from the olfactory bulb traverse the cribriform plate into the nasal mucosa, providing minimally invasive access to the central nervous system (CNS). We hypothesized that nasal fluid from this region could enable detection of neurology-relevant proteins using targeted multiplex assays. Using nosecollect®, a targeted nasal sampling device, nasal fluid proximal to brain-nose interface was collected from cognitively impaired patients, alongside matched cerebrospinal fluid (CSF) and plasma. After nasal sample-specific dilution optimization and intra-assay precision evaluation, all matrices were profiled with the Olink Target 96 Neurology and NUcleic acid Linked Immuno-Sandwich Assay CNS disease 120 (NULISAseq CNS Disease 120) panels. Nasal fluid showed technically repeatable detection (intra-assay coefficient of variation <10% for more than 60% of proteins). Target detectability in nasal fluid (Olink 89/92; NULISA 121/131 proteins) was comparable to plasma and exceeded CSF. Numerous disease-relevant proteins were observed in nasal fluid, including brain-derived tau species, phosphorylated tau, α-synuclein, axonal and synaptic markers, and glial-microglial mediators. To our knowledge, this is the first neurology-focused characterization of the nasal proteome from the vicinity of brain-nose interface using targeted multiplex platforms and the first to profile matched nasal fluid, CSF, and plasma, supporting nasal fluid from brain-nose interface as an additional source for biomarker development.

## Introduction

Neurodegenerative disorders have long preclinical phases in which pathology accumulates before symptoms such as cognitive decline become apparent. Fluid biomarkers in cerebrospinal fluid (CSF) and blood now allow diagnosis and staging of these processes. CSF biomarkers are incorporated into diagnostic criteria, and ultrasensitive assays have enabled measurement of many of these proteins in plasma, expanding clinical and research applications^1–3^. Yet, CSF collection via lumbar puncture is invasive and poorly suited to frequent sampling, while blood biomarkers integrate central and peripheral signals and can be confounded by systemic comorbidities and peripheral metabolism^4,5^. Therefore, there is a persistent need for complementary, minimally invasive biospecimens that yield central nervous system (CNS)-relevant molecular information and are suitable for applications that require repeated sampling such as disease or treatment response monitoring.

The brain-nose interface (BNI) represents a unique and underexplored access point to CNS-related molecular signals. At the level of the olfactory mucosa, millions of olfactory receptor neurons project through the cribriform plate to synapse in the olfactory bulb, establishing direct anatomical continuity between intra- and extracranial compartments (Fig. 1). This continuity across the cribriform plate has been implicated in the efflux of CSF from the brain toward the nasal cavity^6,7^. Experimental tracer studies in multiple species, ranging from rodents to humans, have shown that tracers injected intracranially to CSF compartment accumulates along olfactory nerve bundles, traverses the cribriform plate, and enters lymphatic vessels in the nasal mucosa and downstream cervical lymph nodes, indicating a substantial CSF outflow route via the BNI^7–10^. Together, these observations raise the possibility that nasal fluid collected directly at the BNI could harbour CNS-derived proteins relevant to neurodegeneration. Supporting this, multiple post-mortem, *in vivo*, and biomarker studies have shown that biomarker proteins such as Amyloid-β (Aβ), Tau, α-synuclein can be detected in olfactory mucosa and nasal samples across Alzheimer’s disease (AD) and other neurodegenerative diseases using immunoassays and seed-amplification assays^11–14^.

**Figure 1:**
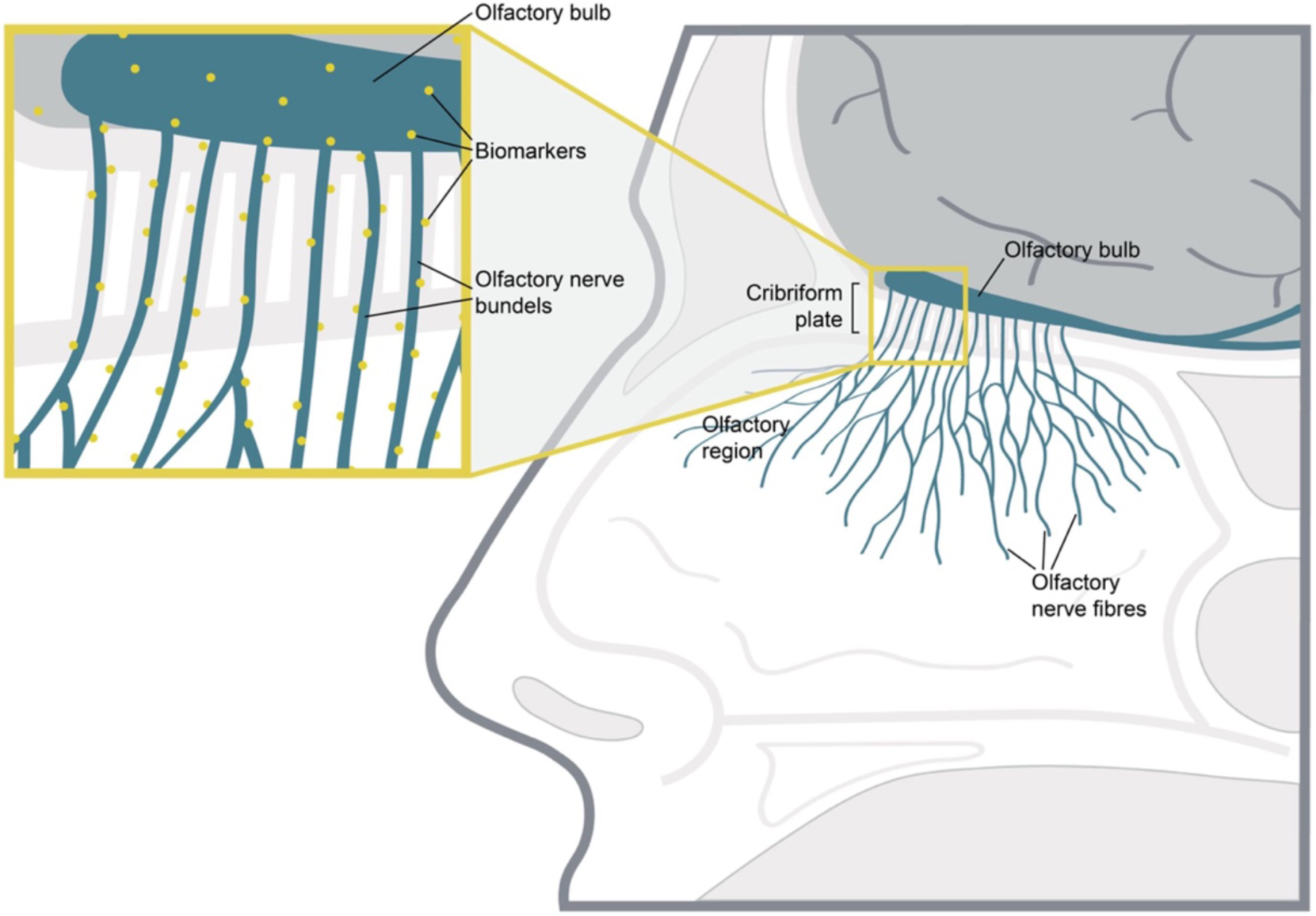
Schematic overview of the BNI. The BNI represents the anatomical junction between the nasal olfactory mucosa and the olfactory bulb, with olfactory nerve fibers traversing the cribriform plate and connecting the intra- and extra- cranial regions. This direct anatomical continuity provides a conduit for molecular exchange from brain to nose (Reproduced from San Nicoló, Marion, et al. “Novel, standardized sample collection from the brain-nose interface.” Methods 234 (2025): 233-241.,^30^ under the terms of the Creative Commons Attribution 4.0 International License (CC BY 4.0) (http://creativecommons.org/licenses/by/4.0/)).

To date, neurology-focused biomarker studies in nasal fluids have mostly focused on single or few analytes and were based on sampling strategies not specifically optimized for targeted sample collection from the BNI^11–13,15^. Nasal sample studies prominently utilised methods such as nasal lavage, nasal swabs, brushes, or by generic absorptive methods placed on the nasal mucosa (eg: nasosorption) for sample collection, none of which were designed to selectively access exclusively the upper nasal cavity in the vicinity of the BNI region^11,12,16,17^. In parallel, untargeted mass spectrometry-based proteomics has been applied to nasal samples in other diseases ((e.g., chronic rhinosinusitis, allergic rhinitis)), identifying hundreds to thousands of proteins and defining inflammatory, epithelial, and immune signatures, but not using anatomically targeted BNI sampling or focusing on individuals with cognitive impairment or at risk for neurodegenerative disease^18–24^. Overall, neurology-focused, multiplex investigations of BNI-derived nasal samples remain scarce, and most prior work has not systematically compared nasal fluid biomarker profiles with matched CSF and plasma. Consequently, the overall neurology-relevant proteomic landscape of nasal fluids at the BNI in individuals at risk for CNS disorders remains poorly characterized.

Recent advances in targeted proteomics, including proximity extension assays like Olink and ultrasensitive immunoassay platforms such as Nucleic acid–linked immuno-sandwich assay (NULISA), now enable simultaneous quantification of tens to hundreds of low-abundance proteins in small-volume biofluids (e.g., plasma, CSF)^25,26^. Olink panels have already been applied to nasal samples in non-neurological settings, demonstrating feasibility of high-plex proximity assays in this matrix, but without anatomically targeted BNI sampling or a CNS focus^16,17,27–29^. Similarly, to date no studies have evaluated a CNS-focused NULISA panel in nasal samples. Applying these neurology-focused platforms to BNI-derived nasal fluid offers an opportunity to investigate the feasibility of detecting neurology-and CNS-related proteins in this matrix and assess how nasal profiles relate to those in CSF and plasma in cognitively impaired individuals. To enable such BNI-focused nasal sampling, we have previously developed and described a minimally invasive device called nosecollect®, that collects nasal fluid from the BNI region in a targeted manner for downstream analysis^30^.

In this study, we performed parallel Olink Target 96 Neurology and Nucleic acid-Linked Immuno-Sandwich Assay paired with Next-Generation Sequencing (NULISAseq) CNS disease 120 (also referred to as NULISAseq CNS for brevity) profiling in nasal fluid obtained at the BNI, as well as in matched CSF and plasma, from patients with cognitive impairment. Nasal fluid from the BNI was collected with nosecollect®. We evaluated the feasibility of CNS/neurology-focused targeted multiple proteomics in nasal fluid by characterizing neurology-related protein detectability across these three matrices. We also quantified cross-matrix overlap and abundance correlations. By integrating high-content proteomics with anatomically targeted sampling at the BNI and direct comparison to established fluid biomarker matrices, this work provides a foundational characterization of the CNS-relevant and neurodegeneration associated BNI proximal nasal proteome.

## Materials and Methods

### Study Cohort and ethics approval statement

As CSF sampling from healthy individuals is not easily justifiable due to ethical concerns, a cognitively impaired cohort was chosen due to the availability of CSF that is routinely collected for their assessment. All clinical samples used in this study originated from patients with cognitive impairment recruited at affiliated memory clinics (Charité - Universitätsmedizin Berlin, Friedrich-Alexander-Universität (FAU) Erlangen, Universitätsmedizin Göttingen, Zentralinstitut für Seelische Gesundheit, Mannheim). This study involving human participants was conducted in accordance with the principles of the Declaration of Helsinki and followed applicable laws and ethical guidelines. Participants were enrolled in the clinical study with Ethics Committee approval by Ethik-Kommission der Bayerischen Landesärztekammer (approval number 21112; date of approval: 03-FEB-2022), and the study has been registered on ClinicalTrials.gov (ClinicalTrials.gov identifier: NCT05791552). Participants provided informed consent to participation prior to inclusion. Participants’ privacy rights were respected, and all data were handled in accordance with relevant data protection and confidentiality regulations. The nasal fluid, plasma and CSF samples were collected over a period ranging from April 2023 to September 2024. After collection at the respective centres, samples were processed according to the study protocol (described in the following section) and subsequently stored at Noselab GmbH at −80 °C until use.

### Sample Collection and Handling and Transport

Nasal fluid from the BNI region was collected using the nosecollect® device (developed by Noselab GmbH) that positions an absorption material in the upper nasal cavity in close proximity to the BNI area in a minimally invasive manner on both sides. The absorbent element is enclosed within the distal tip of the device during insertion and is deployed in a targeted manner at the time of sampling. The use of nosecollect® for BNI-directed nasal fluid collection has been previously evaluated and reported by San Nicolo et al., 2025^30^.

Nasal fluid collection with nosecollect® was performed in the clinic by medical staff trained in the use of the device. After collection, the absorption materials from the BNI from both sides of the nose were stored in an Eppendorf Protein LoBind® transportation tube. CSF samples were collected via lumbar puncture according to standard clinical procedures, and blood samples were obtained by routine venipuncture as specified by the local clinical laboratory protocol.

Immediately after sample collection at the clinics, all three sample types (absorption material in transport tubes, CSF, and plasma) were frozen and stored at -80° until transport. Samples were shipped on dry ice to the biobank facility at Noselab GmbH, where nasal fluid was eluted, and all samples were stored at -80 °C until further use. For the Olink and NULISA measurements, samples were later shipped on dry ice from the Noselab GmbH to the respective measurement sites. Olink assays were performed at the Helmholtz Munich- Core Facility Metabolomics and Proteomics, and NULISA experiments were conducted at the Deutsches Rheuma-Forschungszentrum (DRFZ, Berlin) and at the Platform for Genomics and Epigenomics (PRECISE) at the German Center for Neurodegenerative Diseases (DZNE) in Bonn. Upon arrival, samples were stored at −80 °C at the measurement sites until the day of analysis.

### Olink experimental protocol

Olink experiments were performed at the Helmholtz Munich- Core Facility Metabolomics and Proteomics. Before performing Olink experiments with matched samples, we assessed dilution-dependent detectability and technical repeatability of the Olink Target 96 Neurology panel using nasal fluid samples. In the dilution-optimization experiment, 6 nasal fluid samples from cognitively impaired subjects were analyzed. Nasal fluid samples were measured undiluted and at 1:4, 1:8, 1:16, and 1:32 dilutions prepared in either Olink Predilution Buffer + 1× cOmplete Mini protease inhibitor (Roche #11836153001) or 1× Radioimmunoprecipitation assay (RIPA) lysis buffer (Millipore #20-188) + 1× protease inhibitor. Three of the nasal fluid samples were run in duplicate across all buffer-dilution conditions, while the other three samples were run as single measurements.

In the matched-sample experiment, 48 nasal fluid, 24 CSF and 16 plasma samples were analyzed using the same Olink Target 96 Neurology panel. Olink provided Normalized Protein Expression (NPX) values, a log_2_-scaled relative abundance measure.

Olink optimization experiments were performed in March 2025 and matched sample measurements were performed in May 2025.

### NULISA experimental protocol

Before performing NULISA experiments with matched samples, we assessed dilution-dependent detectability of the NULISA using nasal fluid samples. For dilution optimisation of nasal fluid on NULISA, nasal fluid samples were analyzed using the NULISAseq Inflammation Panel 250. For dilution optimization, two nasal fluid samples from cognitively impaired subjects were measured undiluted and diluted in phosphate-buffered saline (PBS) (1:4, 1:8, 1:16, 1:32). The resulting dilution curves were used to select the working dilution for subsequent measurements. These dilution optimisation experiments were performed at Deutsches Rheuma-Forschungszentrum (DRFZ, Berlin) in February 2025.

After establishing the working dilution, matched samples (47 nasal fluid, 22 CSF, 16 plasma) were analyzed using the 131-plex NULISAseq CNS Disease 120 Panel. One nasal fluid sample was run in duplicate and used to estimate technical variability across wells. These experiments were performed at PRECISE (DZNE, Bonn) in June 2025.

NULISA experiments were performed following the standard NULISAseq workflows for the respective panels using an ARGO HT instrument and followed by sequencing on an Illumina NextSeq 2000 system with a NextSeq™ 1000/2000 P2 XLEAP-SBS™ Reagent Kit (100 Cycles). The raw Illumina BCL files were demultiplexed with bcl2fastq2. The FASTQ files were processed using NULISA Analysis Software to obtain per-target read counts and normalized protein quantities on a log₂ scale called Normalized Protein Quantification (NPQ).

### Statistical analysis

#### Data preprocessing

Log₂-scale NPX (Olink) and NPQ (NULISA) values were used as provided for downstream analysis. All sample-level and assay-level quality control criteria were met for Olink, in both dilution optimisation and matched sample experiments. While all run-level quality control criteria were met for NULISA in both dilution optimisation and matched sample experiments, a subset of samples in the matched experiment had low read count warnings for CSF and plasma (CSF=5/22, plasma=14/16). Additionally, three targets (CRH, NPTX1, and pTau-217) exhibited intra-plate coefficients of variation (CV) >30% based on platform-defined sample controls (SC; pooled plasma replicates run on each plate as an independent measure of technical reproducibility). While all samples and targets were retained for downstream analysis, only measurements above the platform-specific limit of detection (LOD) were included, unless otherwise specified. Proteins were only classified as detectable within a given matrix type if measurements were above LOD in ≥50% of samples.

#### Technical variability

When duplicate measurements were available, technical variability was assessed using intraclass correlation coefficients (ICC), CV, and Bland-Altman analysis. ICC was calculated using a two-way random-effects, absolute-agreement model for single measurements (ICC(2,1)), which partitions total variance into between-protein differences and measurement error across technical replicates. CV was computed on back-transformed linear-scale values as the standard deviation divided by the mean across technical duplicates. For Olink, NPX values were back-transformed as 2^NPX^; for NULISA, NPQ values were back-transformed as 2^NPQ^−1 prior to CV calculation. Bland-Altman analysis was performed on log₂-scale values (NPX or NPQ) to estimate systematic bias and limits of agreement between technical replicates, defined as the mean difference ± 1.96 standard deviations.

CV distributions across proteins were visualized to assess repeatability. For Olink, where three samples were run in duplicate, empirical cumulative distribution function (ECDF) curves were used to compare distributions across samples. For NULISA, where only a single duplicate sample was available, a histogram was used instead.

#### Detectability, overlap, and cross-matrix concordance

Detection rates were summarized as the proportion of samples in which a protein was measured above LOD. For each matrix, proteins measured above LOD in ≥50% of samples were considered as reliably detected. Overlap across matrices was visualized using Venn diagrams. Cross-matrix concordance was assessed using Spearman rank correlation coefficients computed protein-wise across matched samples. Correlations were calculated using only measurements above LOD in both matrices, and only those proteins were included in correlation analyses for which at least three paired observations above LOD were available in both sample types.

#### Concordance across Olink and NULISA

Cross-platform concordance was evaluated for analytes measured on both Olink Target 96 Neurology and NULISAseq CNS panels. Overlapping targets across the two panels were identified based on matching UniProt identifiers. For each overlapping protein, paired measurements across matched samples were compared within each matrix (nasal fluid, CSF, plasma). Analyses were performed on log₂-scale values (NPX for Olink and NPQ for NULISA). The number of matched samples across Olink Target 96 Neurology and NULISAseq CNS panels per matrix was n = 47 (nasal fluid), n = 22 (CSF), and n = 16 (plasma).

Spearman rank correlation coefficients (ρ) were computed to quantify monotonic agreement between platforms. For each analyte, correlations were calculated using only paired samples in which measurements were above the respective platform-specific LOD in both assays. Proteins were included if at least three such paired observations were available. Statistical significance was assessed using two-sided tests, with p < 0.05 considered significant.

### Software

All analyses were performed in Python (version 3.11) using pandas, scipy, and pingouin. Figures were generated using matplotlib and seaborn.

## Results

### Dilution optimisation and technical variability of nasal fluid on Olink

Before performing experiments with matched samples, we assessed dilution-dependent detectability and technical repeatability of the Olink assay using nasal fluid samples from cognitively impaired subjects (Fig. 2).

**Figure 2:**
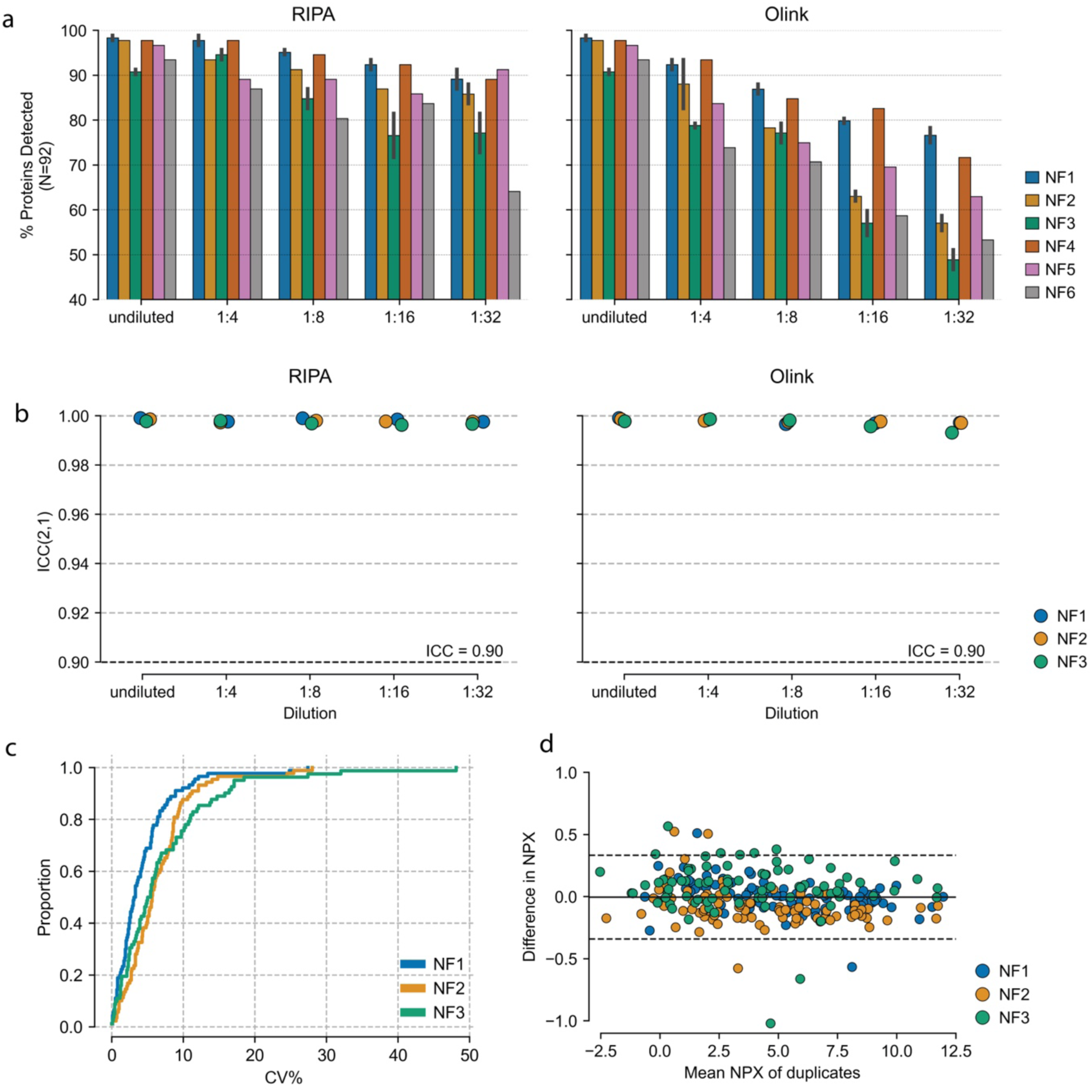
Detection performance and technical variability of protein measurements in nasal fluid (NF) samples on Olink. **(a)** Percentage of proteins detected in multiple nasal fluid samples (NF1-NF6) across undiluted and buffer-diluted conditions (RIPA and Olink buffer). Samples NF1-NF3 were measured in technical duplicates, whereas samples NF4-NF6 were measured as single measurements. **(b)** Intraclass correlation coefficient, ICC (2,1), for the duplicate measurements of NF1-NF3, quantifying within-sample agreement by estimating the proportion of total variance attributable to true signal rather than measurement error; ICC values near 1 indicate high reproducibility. **(c)** Empirical cumulative distribution function (ECDF) of coefficients of variation (CVs) for the duplicate measurements in NF1-NF3, computed on linear-scale expression values (2^NPX^). **(d)** Bland-Altman plots for NF1-NF3, showing the difference in NPX between duplicate measurements against their mean NPX, with the solid line representing the estimated bias and the dashed lines the 95% limits of agreement (±1.96 SD).

To identify the optimal dilution that maximised detectability, dilution series experiments were performed across a range of nasal fluid dilutions (undiluted, 1:4, 1:8, 1:16, 1:32) using six samples per dilution with Olink Target 96 Neurology panel (n=92 proteins). Measurements were performed in both Olink buffer and RIPA buffer to assess the effect of buffer composition on detectability. We found that most proteins (>90%) were detected above LOD in undiluted samples, with a progressive decline in detectability at higher dilution factors irrespective of the buffer type (Fig. 2a). To maximize protein coverage in the matched experiments, we therefore proceeded with undiluted nasal fluid samples for the matched sample experiment on Olink.

Repeatability of nasal fluid measurement on Olink Target 96 Neurology panel was assessed using intraclass correlation coefficients (ICC(2,1)), coefficients of variation (CV), and Bland-Altman analysis and was found to be excellent. Duplicate measurements were available for n=3 cognitively impaired patient samples corresponding to the dilutions 1:4, 1:8, 1:16, 1:32 as well as its undiluted condition. Duplicate measurements (samples NF1-NF3) yielded ICC(2,1) values ≥0.99, with most proteins (NF1 90.2%; NF2 84.8%; NF3 67.4%) exhibiting CVs <10% and only a small fraction (less than 10% across all samples) exceeding 20% CV (Fig. 2b-c). Bland-Altman analysis demonstrated minimal systematic bias and narrow limits of agreement (Fig. 2d).

### Dilution optimisation and technical variability of nasal fluid on NULISA

Similar to Olink, we also evaluated the dilution-dependent detectability and analytical performance of NULISA assay in nasal fluid samples collected from cognitively impaired subjects (Fig. 3).

**Figure 3:**
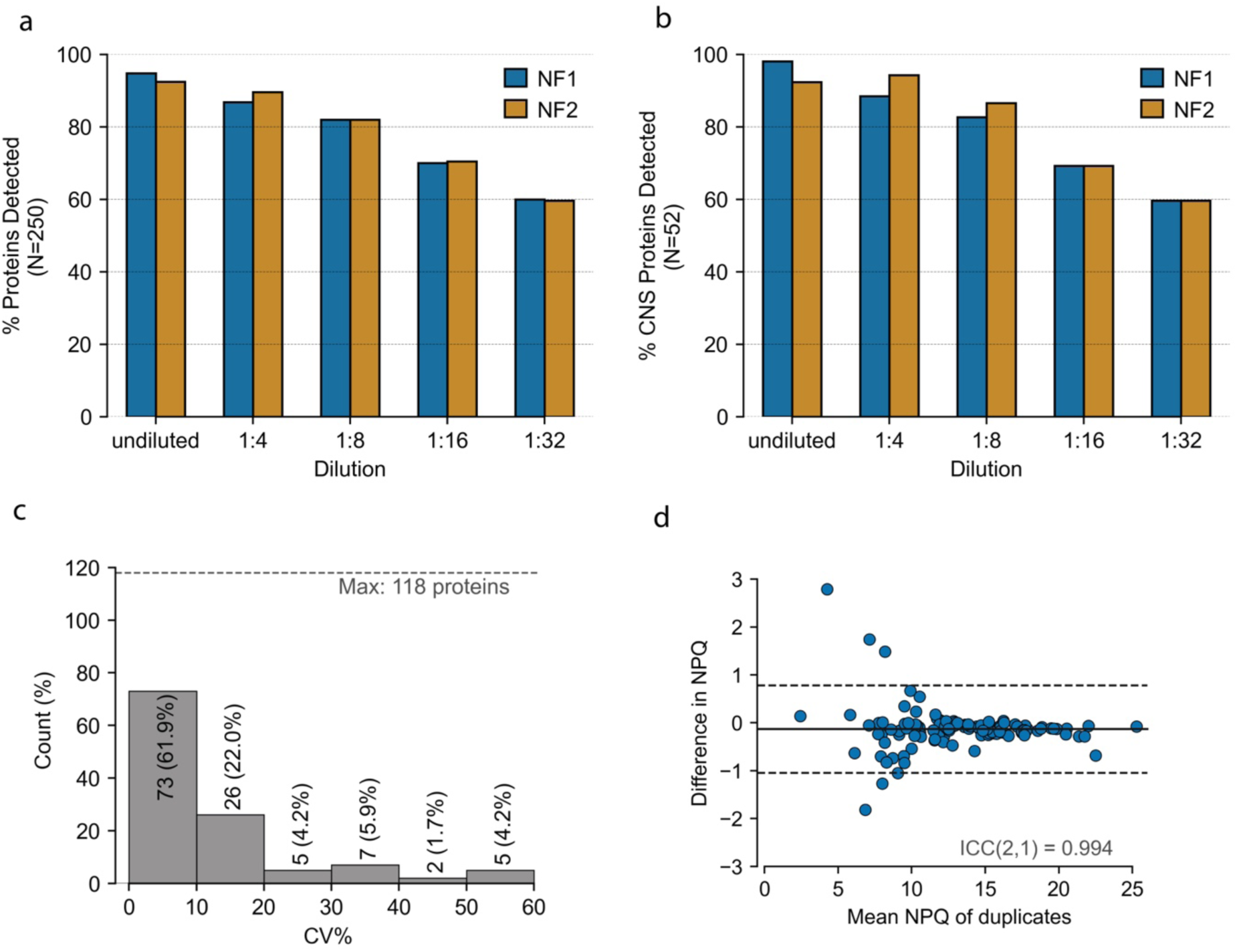
Detection performance and technical variability of protein measurement in nasal fluid (NF) samples on NULISA. **(a)** The percentage of NULISAseq Inflammation panel proteins detected in two nasal samples across serial dilutions (undiluted, 1:4, 1:8, 1:16, 1:32). **(b)** The percentage of NULISAseq Inflammation panel proteins that are also present on the CNS panel (52 overlapping proteins) detected in the same two nasal samples across dilution series. **(c)** Distribution of coefficients of variation (CVs, %) across proteins based on duplicate measurement of a nasal fluid sample (computed on linear-scale NPQ values). Majority of proteins exhibit low technical variability (CV <10%). The dashed horizontal line indicates the total number of proteins evaluated (n = 118; both duplicates above LOD). **(d)** Bland-Altman plot of duplicate measurements showing the difference in NPQ versus the mean NPQ for each protein. The solid line represents the estimated bias, and the dashed lines indicate the 95% limits of agreement (±1.96 SD). Intraclass correlation (ICC(2,1) = 0.994) indicates excellent repeatability.

Dilution optimization was performed using the NULISAseq Inflammation panel (n=250 proteins) across a range of dilutions (undiluted, 1:4, 1:8, 1:16, 1:32) using two samples per dilution. Similar to Olink, nasal fluid samples showed a comparable dilution-dependent decline in detectability, decreasing from ∼93% (undiluted) to ∼60% (1:32) of targets (Fig. 3a). To directly compare the effect of dilution observed in inflammation panel with the NULISAseq CNS panel used for the matched experiments, we analysed the effect of dilution on the 52 targets overlapping between the CNS and Inflammation panels (Fig. 3b). Within this overlapping subset, undiluted samples consistently achieved the highest detection rates, supporting the use of undiluted nasal fluid samples in the matched experiments with NULISAseq CNS panel.

For NULISA, CVs were calculated using duplicate measurements of a nasal fluid sample from a cognitively impaired patient measured with the NULISAseq CNS panel (131 proteins). Only proteins with both measurements above LOD were included (n=118). Similar to Olink, NULISA duplicates exhibited predominantly low variability, with 61.9% of proteins showing CV <10%, only 16.1% exceeding 20%, and an ICC(2,1) of 0.994, indicating close agreement between replicates (Fig. 3c,d)

Together, these analyses confirm the technical suitability of nasal fluid for high-plex proteomic profiling, forming the basis for the matched cross-matrix experiment described below.

### Protein detectability using Olink and NULISA

Having established the conditions for analysis of nasal fluid, we proceeded to profile matched nasal fluid, CSF, and plasma samples across both Olink and NULISA platforms. The samples were acquired from cognitively impaired subjects at memory clinics (Fig. 4a). While samples were collected from the same cohort, not all participants contributed all three matrices, resulting in a partially matched design that enables within-subject comparisons where data are available. Nasal fluid was available from 48 individuals, with Olink data for all 48 and NULISA data for 47 (Table 1). From the same cohort, CSF was analyzed in 24 individuals with Olink and 22 with NULISA (Table 2), and plasma in 16 individuals with both platforms (Table 3). Cohort characteristics corresponding to the three matrices are summarised in Table 1–3 and SI Tables 1–3.

**Figure 4:**
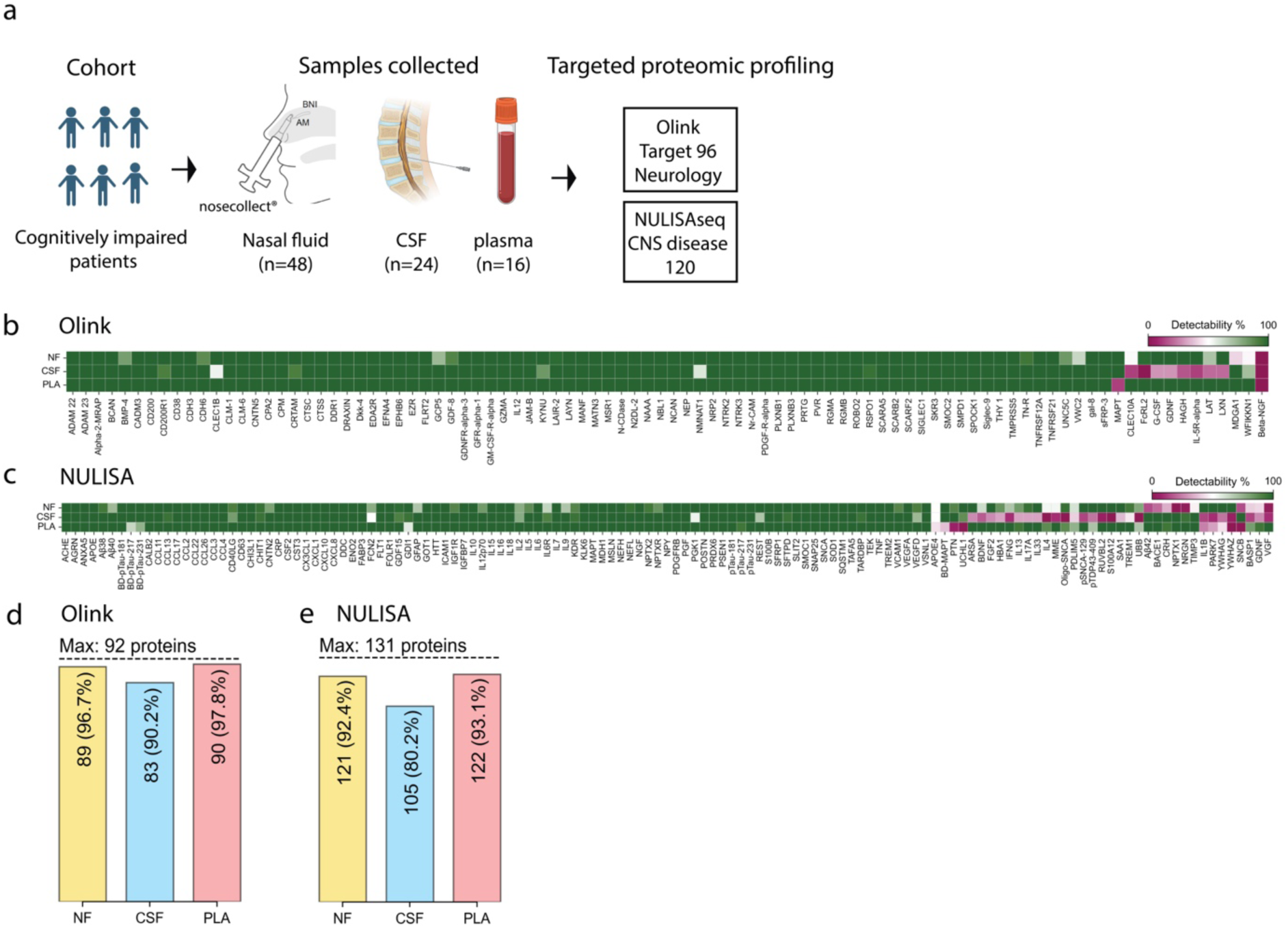
Protein detectability profiles for Olink and NULISA platforms across sample types. **(a)** Study workflow: The study included 48 patients with mild cognitive impairment. Nasal fluid samples were obtained from all 48 patients while CSF and plasma samples were available for 24 and 16 of these patients, respectively. The matched samples were profiled using the Olink Target 96 Neurology panel and the NULISAseq CNS panel. All available samples were included in the Olink Target 96 Neurology panel measurements. From the available samples, NULISAseq CNS analyses lacked one nasal fluid sample and two CSF samples. Created in BioRender (https://BioRender.com). (Adapted in part from San Nicoló, Marion, et al. “Novel, standardized sample collection from the brain-nose interface.” Methods 234 (2025): 233-241.,^30^ under the terms of the Creative Commons Attribution 4.0 International License (CC BY 4.0) (http://creativecommons.org/licenses/by/4.0/)) **(b, c)** Heatmap of protein detectability (0-100%) for Olink (b) and NULISA (c) across nasal fluid (NF), CSF and plasma (PLA). Each row in the heatmaps correspond to a specific sample type and each column corresponds to an individual protein target (labels along the x-axis). Color corresponds to the percentage of samples in which that protein was detected (i.e., above LOD), with green indicating higher detectability and purple lower. (**d, e**) The overall performance of the Olink (d) and NULISA (e) panels, respectively, showing the total number and proportion of proteins with at least 50% detectability per matrix.

**Table 1:** Cohort characteristics of nasal fluid samples used for Olink and NULISA experiments.

| Methodology | Olink | NULISA |
| --- | --- | --- |
| Total n | 48 | 47 |
| Gender:female n (%) | 24 (50%) | 24 (51.1%) |
| Age mean ± SD (median, min-max) | 70.5 ± 8.5 (73.0, 52.0-82.0) | 70.8 ± 8.3 (73.0, 52.0-82.0) |
| MMSE mean ± SD (median, min-max) | 25.4 ± 3.9 (26.0, 10.0-30.0) | 25.4 ± 4.0 (26.0, 10.0-30.0) |
| MoCA mean ± SD (median, min-max) | 20.2 ± 5.5 (20.5, 3.0-28.0) | 20.1 ± 5.6 (20.0, 3.0-28.0) |

**Table 2:**
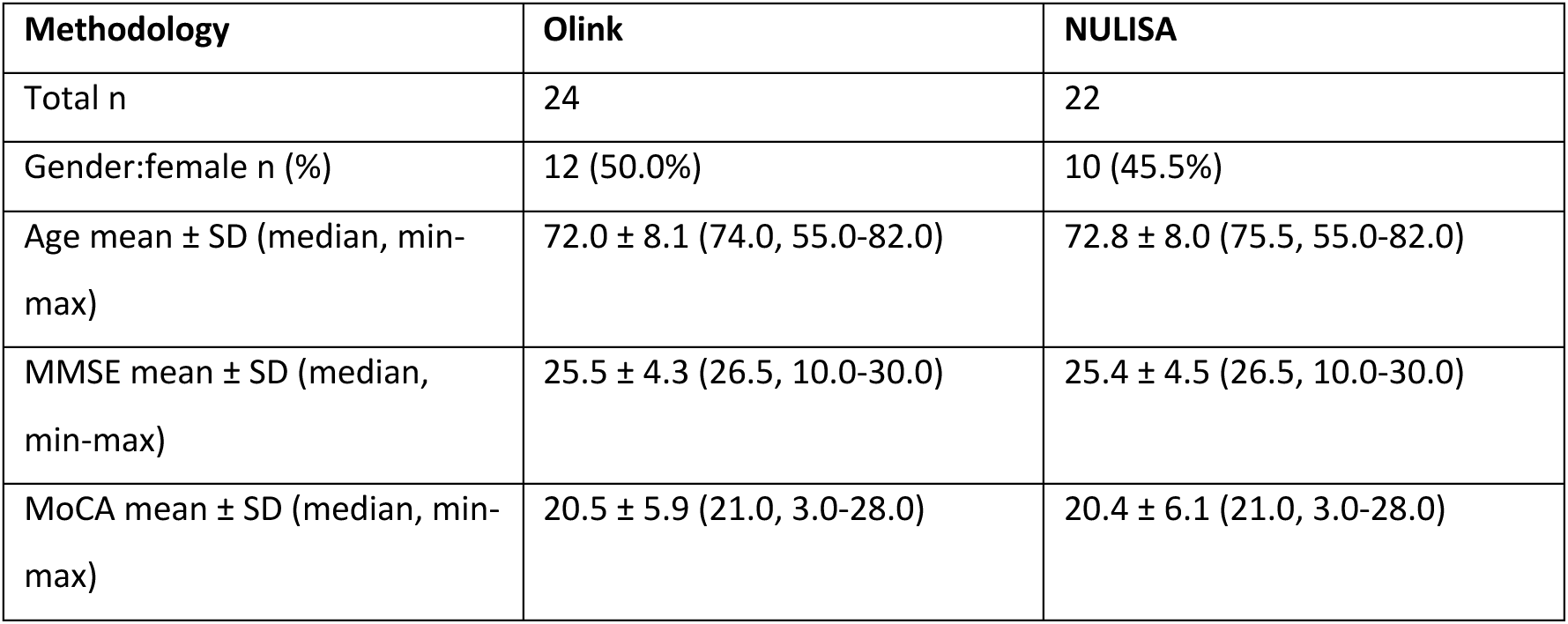
Cohort characteristics of CSF samples used for Olink and NULISA experiments.

| Methodology | Olink | NULISA |
| --- | --- | --- |
| Total n | 24 | 22 |
| Gender:female n (%) | 12 (50.0%) | 10 (45.5%) |
| Age mean ± SD (median, min-max) | 72.0 ± 8.1 (74.0, 55.0-82.0) | 72.8 ± 8.0 (75.5, 55.0-82.0) |
| MMSE mean ± SD (median, min-max) | 25.5 ± 4.3 (26.5, 10.0-30.0) | 25.4 ± 4.5 (26.5, 10.0-30.0) |
| MoCA mean ± SD (median, min-max) | 20.5 ± 5.9 (21.0, 3.0-28.0) | 20.4 ± 6.1 (21.0, 3.0-28.0) |

**Table 3:** Cohort characteristics of plasma samples used for Olink and NULISA experiments.

| Methodology | Olink | NULISA |
| --- | --- | --- |
| Total n | 16 | 16 |
| Gender:female n(%) | 8 (50.0%) | 8 (50.0%) |
| Age mean ± SD (median, min-max) | 72.3 ± 8.7 (75.5, 55.0-82.0) | 72.3 ± 8.7 (75.5, 55.0-82.0) |
| MMSE mean ± SD (median, min-max) | 25.4 ± 5.1 (27.5, 10.0-30.0) | 25.4 ± 5.1 (27.5, 10.0-30.0) |
| MoCA mean ± SD (median, min-max) | 20.8 ± 6.7 (22.5, 3.0-28.0) | 20.8 ± 6.7 (22.5, 3.0-28.0) |

We first characterized protein detectability across matrices using the Olink Target 96 Neurology and NULISAseq CNS panels (Fig. 4). Proteins for which at least 50% of measurements were above platform-specific LOD were considered to be reliably detected. Nasal fluid exhibited consistently high detectability across both platforms (Fig. 4b-e, SI Table 2,3): 89 out of 92 proteins (96.7%) were detected by Olink Target 96 Neurology panel and 121 proteins out of 131 (92.4%) were detected by NULISAseq CNS panel. These values were comparable to those observed in plasma and higher than that of CSF. Olink detected up to 90 proteins in plasma (97.8%) and 83 in CSF (90.2%), while 122 (93.1%) and 105 (80.2%) proteins were detected in plasma and CSF, respectively, using NULISA.

### Cross-matrix comparison of proteomes detected by NULISAseq CNS Disease 120 and Olink Target 96 Neurology panels across nasal fluid, CSF, and plasma

On both Olink Neurology and NULISAseq CNS panels, the majority of neurology-focused targets formed a shared core proteome that was detected in nasal fluid, CSF, and plasma (Fig. 4b,c and Fig. 5a,b). This core included many neurodegenerative disease associated proteins (MAPT, PSEN1, SNCA/total α-synuclein, Aβ40, APOE)^2,31–33^, axonal damage related proteins(eg: NEFL, NEFH^34^), synaptic dysfunction related proteins(eg: SNAP25, NPTX2, NPTXR)^35,36^, and glial activation/neuroinflammation related proteins (eg: CHI3L1, S100B, TREM2^37^), all of which were detected in more than 50% of samples across nasal fluid, CSF, and plasma. However, beyond this core, the heatmaps also highlight several non-overlapping subsets that distinguish the three fluids (Fig. 4b,c, Fig. 5a,b, and SI Tables 2,3).

**Figure 5:**
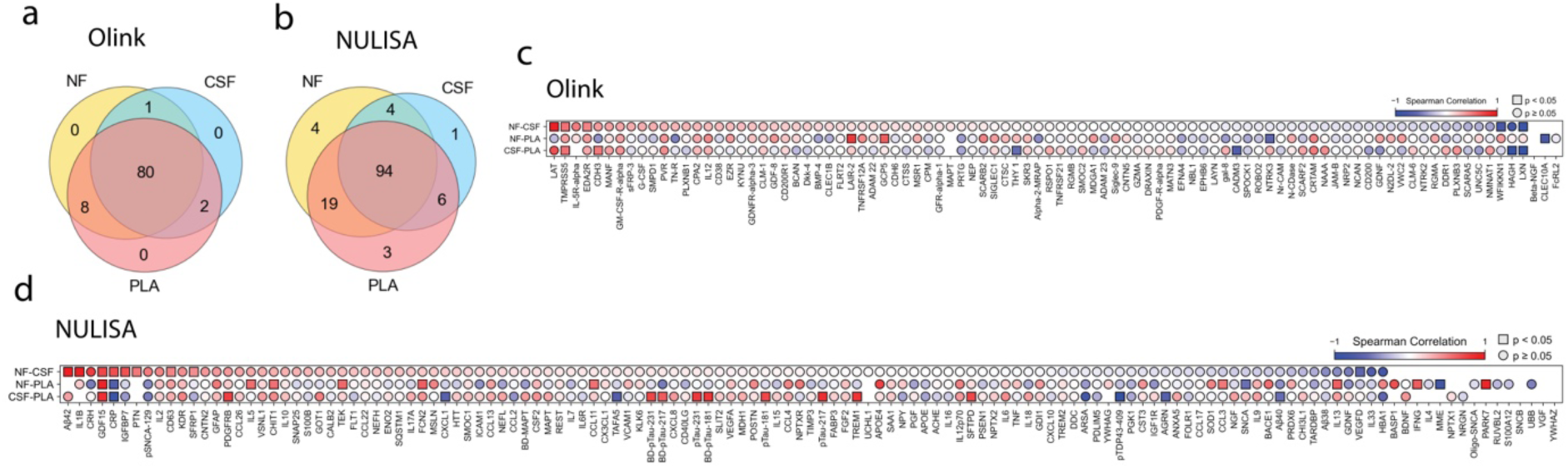
Overlap and correlations of protein measurements on Olink and NULISA platforms. **(a,b)** Venn diagrams summarizing the overlap in detectable proteins (above LOD in ≥50% of samples) across nasal fluid (NF), CSF and plasma (PLA) for Olink (a) and NULISA (b). **(c,d)** Spearman correlations of protein levels between nasal fluid (NF), CSF and plasma (PLA) (marked as NF-CSF, NF-PLA, CSF-PLA) for Olink (c) and NULISA (d). Each column corresponds to a protein, rows indicate matrix pairs, and colors encode the correlation coefficient (blue to red scale from −1 to 1). Squares indicate statistically significant correlations with p < 0.05, while circles represent non-significant correlations.

Interestingly, brain-derived (BD)-MAPT was readily detected in nasal fluid and CSF but fell below the 50% detectability threshold in plasma (nasal fluid 100%, CSF 100%, plasma 38% of samples; Fig. 4c). Similarly, phosphorylated BD-Tau species (BD-pTau-217, BD-pTau-231) showed relatively reduced detectability in plasma compared to nasal fluid and CSF (BD-pTau-217: plasma 62%, nasal fluid: 100%, CSF: 100%; BD-pTau-231: plasma 75%, nasal fluid 100%, CSF 100%; Fig. 4c). In contrast, total Tau (MAPT) and pTau assays (pTau-181, pTau-217, pTau-231) on NULISA showed robust detectability across all three matrices and remained part of the pan-fluid core. (Fig. 4c). As opposed to NULISA, MAPT was not observably detected in plasma on Olink Neurology, and this discrepancy is therefore most likely technical, reflecting lower effective sensitivity and/or epitope choice of the Olink assay (Fig. 4b,c).

In contrast, CSF and plasma also contained a small set of targets with preferential detectability relative to nasal fluid (Aβ42, BACE1, CRH, NPTX1, NRGN, TIMP3, MDGA1, WFIKKN1). (Fig. 4b,c). Notably, Aβ42 was consistently detected in CSF and plasma but only in a subset of nasal fluid samples (nasal fluid 15% vs. CSF and plasma 100% of samples). This is in contrast to other amyloid-pathway components (Aβ38, Aβ40, APOE, PSEN1), which were part of the shared core. Proteins that were predominantly confined to CSF were rare and essentially limited to SNCB.

The detectability analysis also highlighted a broad group of proteins that were abundant in nasal fluid as well as plasma but showed low levels in CSF (Fig. 4b,c). Among this group were key CNS-linked disease-related species, notably Oligo-SNCA, pSNCA-129, and pTDP43-409^38–40^, as well as a prominent subset of inflammatory markers (IL-13, IL-17A, IL-33, S100A12, SAA1, G-CSF, IL-5R-α)^41–44^. Although only few proteins were uniquely confined to nasal fluid or plasma, these still encompassed notable CNS disease-associated proteins, including PARK7 and the 14-3-3 isoforms YWHAZ and YWHAG in nasal fluid, and VGF in plasma (Fig. 4b,c)^45–48^.

### Cross-matrix overlap and correlation between nasal fluid, CSF and plasma

We next examined quantitative concordance for the overlapping proteins across matrices, within each platform (Fig. 5). A substantial proportion of detected proteins were shared across nasal fluid, CSF, and plasma, with 80 proteins (87%) for Olink and 94 proteins (72%) for NULISA detected in ≥50% of samples in all three matrices (Fig. 5a,b). Only small matrix-specific subsets were observed, indicating broadly shared proteome coverage across the three matrices. We then evaluated concordance quantitatively by computing Spearman correlations between protein abundances across nasal fluid, CSF, and plasma (Fig. 5c,d). Quantitative concordance was generally modest, with correlations centered around zero for the majority of proteins on both platforms, suggesting that protein abundance levels are largely matrix-specific.

### Cross-platform concordance between Olink and NULISA

To assess analytical agreement between platforms, we compared overlapping targets between Olink Target 96 Neurology and NULISAseq CNS panels across nasal fluid, CSF, and plasma (Fig. 6). The overlapping subset was identified using UniProt accession identifiers and included Tau species, BDNF, beta-NGF (measured as NGF on NULISA), IL12 (measured as IL12p70 on NULISA) and NEP (encoded by MME on NULISA). On Olink, Tau was quantified as a single total MAPT measure, whereas NULISA provided a panel of distinct Tau readouts, including MAPT and three phosphorylated species (pTau181, pTau217, pTau231), as well as BD-MAPT and the corresponding BD-pTau181, BD-pTau217, and BD-pTau231 measures. We therefore calculated separate correlations between each NULISA Tau measure and Olink MAPT. Spearman correlations were only calculated when ≥3 values were above the LOD in both platforms.

**Figure 6:**
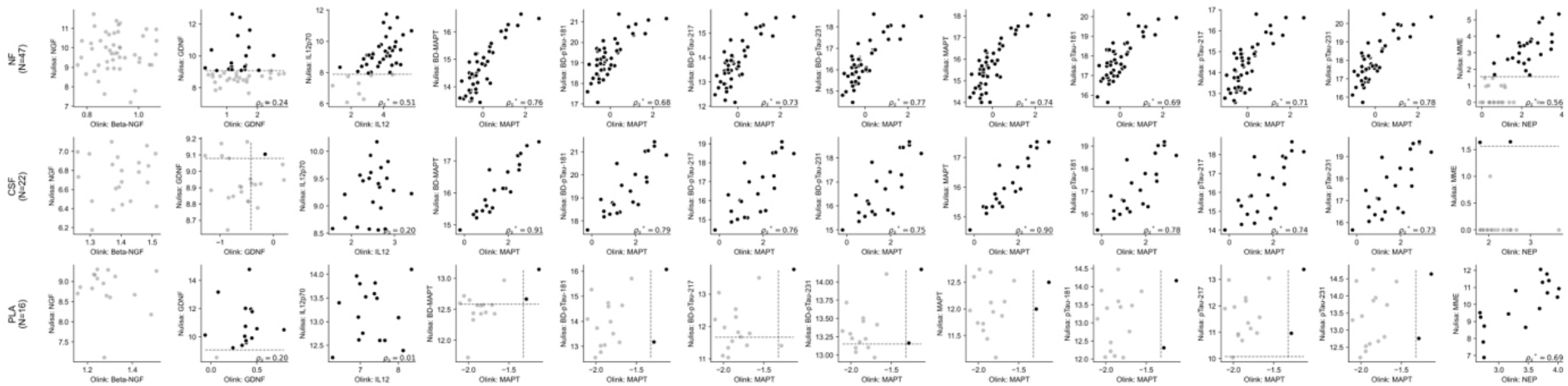
Comparison of Olink vs. NULISA measurements across sample types. Each row represents a different sample type: Nasal fluid (NF) (top), CSF (middle), and plasma (PLA) (bottom). Each scatter plot displays the Olink measurement on the x-axis versus the NULISA measurement on the y-axis. Black dots indicate measurements above the Limit of Detection (LOD), while grey dots represent measurements below LOD; the dashed line indicates the LOD threshold for each assay. Spearman rank correlation coefficients (r_s) are calculated only for samples with ≥3 measurements above LOD in both methods (i.e., at least 3 black dots per panel). P-values < 0.05 are denoted by (*) indicating statistical significance. The correlation values are displayed in each panel.

In nasal fluid (n=47), different Tau species on NULISA showed moderate to strong positive rank correlation with Olink MAPT (ρ ranging from 0.68 to 0.78). Similarly, in CSF (n=22), cross-platform concordance for Tau markers was high across the two platforms, with ρ values between 0.73 and 0.91. On the other hand, for non-tau analytes (β-NGF, GDNF, IL-12/IL-12p70, and NEP/MME), correlations in nasal fluid and CSF were moderate/weak or not estimable because too few samples were above LOD on one or both platforms. In plasma, most measurements for these overlapping analytes fell below the LOD, and thus, a reliable comparison was not possible. Overall, moderate to strong cross-platform concordance was observed for several overlapping tau-related markers including MAPT and multiple pTau isoforms, in both nasal fluid and CSF, while concordance for other analytes was more variable and largely absent in plasma.

## Discussion

In this study, we systematically profiled brain-related proteins in nasal fluid obtained at the BNI, alongside matched CSF and plasma, using two multiplex immunoassay platforms (Olink Target 96 Neurology and NULISAseq CNS disease 120, complemented by NULISAseq Inflammation for optimization). While previous work has demonstrated the feasibility of targeted multiplex proteomics in nasal samples using Olink and other platforms, they were done in healthy individuals or chronic airway disease cohorts and not in CNS-oriented contexts^16,17,27–29^.In those studies, nasal fluid was typically collected either by saline nasal lavage, which dilutes secretions from the entire nasal cavity, or by absorptive strips or sponges positioned on the inferior turbinate or middle meatus. In contrast, our approach uses nosecollect® to sample undiluted nasal fluid specifically from the vicinity of the BNI, providing an anatomically targeted, CNS-proximal matrix. This design has the potential to enhance analyte concentrations for low-abundance neurology markers and improves spatial specificity compared with conventional inferior/middle meatus sampling. Our work demonstrates that nasal fluid derived specifically from BNI using nosecollect® is a suitable matrix for neurology-focused multiplex proteomics in a cognitively impaired cohort. To the best of our knowledge, this represents the first neurology-focused use of these platforms at the BNI.

Our repeatability analyses are in agreement with previous evaluations of biofluids on Olink and NULISA platforms in terms of intra-assay technical variability, as assessed by CVs and ICCs ^49–51^. We directly quantified intra-assay CVs and ICCs in BNI-derived nasal fluid, showing robust technical repeatability for neurology and CNS panel analytes with CVs <10% (>60% of analytes on both platforms) and ICCs ≥0.99 (Fig. 2b,c,d and Fig. 3c,d). This result is also in agreement with prior work in nasal epithelial lining fluid using Olink Target 96 Inflammation, where Zetlen et al. reported that approximately 73% of analytes exhibited intra-assay CVs <10%^17^. In addition to the low technical variability, nasal fluid also exhibited higher or comparable detectability (89/92 Olink Target 96 Neurology and 121/131 NULISAseq CNS proteins) to CSF and plasma. Together, these observations indicate that BNI-derived nasal samples are technically suitable for neurology-focused panel-based protein measurements on Olink and NULISA.

Cross-platform comparisons of overlapping proteins showed concordant signals for several Tau-related analytes across Olink and NULISA in both CSF and nasal fluid, but more variable concordance for other proteins and matrices within this limited overlapping analyte set (Fig. 6). This pattern is consistent with CSF and plasma proteomics studies demonstrating that cross-platform correlations are highly analyte-dependent: only a subset of proteins achieve high concordance between affinity-based platforms, whereas many targets show moderate or low agreement despite being reliably measured^52,53^. For well-established neurodegeneration markers such as NfL, GFAP, and p-tau181, strong correlations across single-plex immunoassays and NULISA assay have been reported in plasma, while markers such as Aβ species exhibit more modest concordance^52^. Such variability is expected, given that different platforms employ distinct capture reagents and epitopes, differ in sensitivity and dynamic range and may preferentially detect different isoforms of a given protein. Overall, these analyte-specific discrepancies underscore the importance of targeted validation when integrating data across proteomic technologies.

The neurology-focused panels showed that disease-associated proteins organize into a large pan-fluid core and several matrix-specific subsets. A prominent CNS-proximal cluster, exemplified by BD-MAPT and related BD-pTau species, was robustly detected in nasal fluid and CSF but attenuated in plasma, consistent with predominant brain origin and efflux along CSF and perineural routes toward the nasal cavity, as suggested by prior imaging and tracer studies of glymphatic and perivascular clearance^54–56^. In contrast, the ubiquitous detection of non-brain derived MAPT and pTau species across all matrices reflects their broader expression in multiple peripheral tissues and thus, higher systemic abundance, rather than CNS efflux^57^. A separate group of targets, such as Aβ42, BACE1, NRGN, and SNCB, showed preferential detectability in CSF with only partial representation in nasal fluid. While Aβ42 has been previously detected in nasal samples, detection levels are highly sensitive to assay format and matrix conditions, and the poor detectability observed in the nasal fluid in our study may reflect epitope masking of aggregation-prone Aβ42 in untreated nasal fluid rather than a complete absence of nasal Aβ42^11,30,58^ .Conversely, several CNS-linked proteins such as Oligo-SNCA, pSNCA-129, pTDP43-409, and PARK7, together with a broad set of inflammatory were better captured in nasal fluid and/or plasma than in CSF. Because Olink and NULISA are targeted panel-based platforms, these patterns necessarily reflect only the neurology- and inflammation-focused portion of the BNI-proximal nasal proteome; our findings therefore motivate future untargeted high-resolution mass spectrometry studies to extend this work toward a more comprehensive, discovery-oriented characterization of the nasal proteomic landscape at the BNI.

This is consistent with nasal fluid representing a distinct biological compartment rather than a peripheral proxy for CSF or plasma. Despite a considerable overlap of reliably detected proteins across three fluids, the relationship of nasal fluid to CSF and plasma is complex and highlights the biological specificity of the BNI compartment. Spearman correlations of protein abundances across matrices were generally modest, and only a subset of proteins showed stronger concordance between nasal fluid, CSF, and plasma. This limited cross-matrix alignment likely reflects matrix-specific biology. The olfactory mucosa constitutes a specialized neuroepithelial and immune niche; proteins in nasal fluid integrate signals from multiple local cell types and CNS-derived components, while local inflammation, regeneration, and environmental exposures can reshape the proteome independently of CSF or blood. CSF, blood, and nasal fluid each have distinct transport and clearance pathways: CSF circulates via ventricular and subarachnoid flow, blood proteins are shaped by systemic metabolism, and nasal fluid is influenced by perineural efflux and mucociliary clearance^6^. This generates different concentration gradients and degradation profiles for the same proteins in each matrix. Even for canonical AD markers such as Aβ and tau, absolute levels and dynamic ranges are known to differ markedly between CSF and blood despite similar disease associations^59–64^. Our findings are consistent with this principle that proteome can be shared across matrices without strong correlation of abundance levels across matrices. At the same time, these biofluid-specific dynamics underscore the importance of developing suitable normalization approaches to extract biologically meaningful and disease-relevant signals.

Taken together, these patterns argue that BNI-derived nasal fluid has a proteome that overlaps extensively with CSF yet also integrates mucosal and systemic inflammatory signals. These observations support viewing BNI-derived nasal fluid not as a simple surrogate for CSF or plasma, but as a complementary readout of a CNS-connected yet biologically distinct compartment. Although this study was not designed to assess diagnostic performance, the proteomic signatures identified here provide a foundation for future biomarker discovery and clinical validation. In summary, these findings support BNI-derived nasal fluid as a minimally invasive and complementary modality to CSF and plasma for studying CNS disorders.

## Data Availability

All data produced in the present study are available upon reasonable request to the authors and completion of a data use agreement.

## Abbreviations

Aβ: Amyloid-β;
AD: Alzheimer’s disease;
BD: Brain-derived;
BNI: Brain-nose interface;
CNS: Central nervous system;
CSF: Cerebrospinal fluid;
CV: Coefficient of variation;
ECDF: empirical cumulative distribution function;
ICC(2,1): Intraclass correlation coefficient;
LOD: Limit of detection;
NPQ: Normalized protein quantity;
NPX: Normalized protein expression;
NULISA: Nucleic acid-Linked Immuno-Sandwich Assay;
NULISAseq: Nucleic acid-Linked Immuno-Sandwich Assay paired with Next-Generation Sequencing;
PBS: phosphate-buffered saline;
PD: Parkinson’s disease;
RIPA: radioimmunoprecipitation assay

## Declaration of generative AI and AI-assisted technologies in the manuscript preparation process

During the preparation of this work the authors used Bomed Advisor AI in order to improve text and identify existing literature. After using this tool/service, the authors reviewed and edited the content as needed and takes full responsibility for the content of the published article.

## Data availability statement

Raw data will be provided by the authors upon request with a data usage agreement.

## Acknowledgements

We would like to thank Charité – Universitätsmedizin Berlin, FAU Erlangen, Universitätsmedizin Göttingen, Zentralinstitut für Seelische Gesundheit, Mannheim for facilitating the participant recruitment and sample collection. We also thank Helmholtz Munich- Core Facility Metabolomics and Proteomics, Deutsches Rheuma-Forschungszentrum (DRFZ, Berlin) and Deutsches Zentrum für Neurodegenerative Erkrankungen (DZNE, Bonn) for providing technical facilities and support for analytical measurements. Additionally, we acknowledge the technical support provided by Nicole Muth at Helmholtz Zentrum Munich- Core Facility Metabolomics and Proteomics We acknowledge the use of Biomed advisor AI for text refinement and BioRender for creation of illustrations (https://BioRender.com).

## Author Contributions

MoB: Formal analysis and visualization, data interpretation and writing original draft, review and editing; MaB: data interpretation and writing original draft, review and editing; GW: conceptualization, methodology; SK: investigation, review and editing; RM: formal analysis; HW: methodology; AP: investigation, data curation; EDD: investigation, data curation; HT: investigation, data curation; KL: investigation, data curation; FH: investigation, data curation; MFM: investigation, data curation; MSN: methodology; MA: methodology, review and editing; SM: conceptualization, methodology, supervision, review and editing.

