## Supplementary Tables 1,2,3 for "Targeted Proteomic Profiling of Nasal Fluid from the Brain-Nose Interface"

**Supplementary File**

**Supplementary Tables**

Table 1: Sample characteristics of nasal fluid, CSF and plasma.

| **Specimen** | **Variable** | **Description** | **Olink** | **NULISA** |
| --- | --- | --- | --- | --- |
| Nasal fluid | Total protein concentration (µg/mL) | mean ± SD  (median, min–max) | 4837.0 ± 2157.3  (4580.0, 1720.0-13200.0) | 4848.0 ± 2179.3  (4750.0, 1720.0-13200.0) |
| Nasal fluid | Initial volume (µL) | mean ± SD  (median, min–max) | 533.8 ± 248.7  (480.0, 145.0-1040.0) | 538.3 ± 249.4  (480.0, 145.0-1040.0) |
| Nasal fluid | Time to freeze (minutes) | mean ± SD  (median, min–max) | 6.0 ± 4.1  (5.0, 1.0-20.0) | 6.0 ± 4.1  (5.0, 1.0-20.0) |
| Nasal fluid | Sample age (days) | mean ± SD  (median, min–max) | 484.3 ± 121.5  (478.0, 230.0-744.0) | 486.4 ± 122.0  (478.0, 230.0-744.0) |
| CSF | Sample age (days) | mean ± SD  (median, min–max) | 409.0 ± 124.1  (365.0, 78.0-568.0) | 397.1 ± 122.5  (365.0, 78.0-568.0) |
| Plasma | Sample age (days) | mean ± SD  (median, min–max) | 329.1 ± 98.0  (365.0, 78.0-365.0) | 329.1 ± 98.0  (365.0, 78.0-365.0) |

Table 2: Detectability values in nasal fluid, CSF and plasma using Olink Neurology panel. For each target, the proportion of samples with detection above LOD is given (1 = 100%).

|  | **Nasal fluid** | **CSF** | **PLA** |
| --- | --- | --- | --- |
| ADAM 22 | 1 | 1 | 1 |
| ADAM 23 | 1 | 1 | 1 |
| Alpha-2-MRAP | 1 | 1 | 1 |
| BCAN | 1 | 1 | 1 |
| BMP-4 | 0.81 | 1 | 1 |
| Beta-NGF | 0 | 0 | 0 |
| CADM3 | 1 | 1 | 1 |
| CD200 | 1 | 1 | 1 |
| CD200R1 | 1 | 0.92 | 1 |
| CD38 | 1 | 1 | 1 |
| CDH3 | 1 | 1 | 1 |
| CDH6 | 0.88 | 1 | 1 |
| CLEC10A | 0.52 | 0.12 | 1 |
| CLEC1B | 1 | 0.54 | 1 |
| CLM-1 | 1 | 1 | 1 |
| CLM-6 | 1 | 1 | 1 |
| CNTN5 | 1 | 1 | 1 |
| CPA2 | 1 | 1 | 1 |
| CPM | 1 | 1 | 1 |
| CRTAM | 1 | 0.92 | 1 |
| CTSC | 1 | 1 | 1 |
| CTSS | 1 | 1 | 1 |
| DDR1 | 1 | 1 | 1 |
| DRAXIN | 1 | 1 | 1 |
| Dkk-4 | 1 | 1 | 1 |
| EDA2R | 1 | 1 | 1 |
| EFNA4 | 1 | 1 | 1 |
| EPHB6 | 1 | 1 | 1 |
| EZR | 1 | 1 | 1 |
| FLRT2 | 1 | 1 | 1 |
| FcRL2 | 0.98 | 0 | 1 |
| G-CSF | 1 | 0.29 | 1 |
| GCP5 | 0.73 | 1 | 1 |
| GDF-8 | 0.9 | 1 | 1 |
| GDNF | 1 | 0.29 | 1 |
| GDNFR-alpha-3 | 1 | 1 | 1 |
| GFR-alpha-1 | 1 | 1 | 1 |
| GM-CSF-R-alpha | 1 | 1 | 1 |
| GZMA | 1 | 1 | 1 |
| HAGH | 1 | 0.17 | 1 |
| IL-5R-alpha | 1 | 0.12 | 1 |
| IL12 | 1 | 1 | 1 |
| JAM-B | 1 | 1 | 1 |
| KYNU | 1 | 0.83 | 1 |
| LAIR-2 | 0.98 | 1 | 1 |
| LAT | 0.69 | 0.21 | 1 |
| LAYN | 1 | 1 | 1 |
| LXN | 1 | 0.21 | 1 |
| MANF | 1 | 1 | 1 |
| MAPT | 1 | 1 | 0.12 |
| MATN3 | 1 | 1 | 1 |
| MDGA1 | 0.42 | 1 | 1 |
| MSR1 | 1 | 1 | 1 |
| N-CDase | 1 | 1 | 1 |
| N2DL-2 | 1 | 1 | 1 |
| NAAA | 1 | 1 | 1 |
| NBL1 | 1 | 1 | 1 |
| NCAN | 1 | 1 | 1 |
| NEP | 1 | 1 | 1 |
| NMNAT1 | 1 | 0.58 | 1 |
| NRP2 | 1 | 1 | 1 |
| NTRK2 | 1 | 1 | 1 |
| NTRK3 | 1 | 1 | 1 |
| Nr-CAM | 1 | 1 | 1 |
| PDGF-R-alpha | 1 | 1 | 1 |
| PLXNB1 | 1 | 1 | 1 |
| PLXNB3 | 1 | 1 | 1 |
| PRTG | 1 | 1 | 1 |
| PVR | 1 | 1 | 1 |
| RGMA | 1 | 1 | 1 |
| RGMB | 1 | 1 | 1 |
| ROBO2 | 1 | 1 | 1 |
| RSPO1 | 1 | 0.96 | 1 |
| SCARA5 | 1 | 1 | 1 |
| SCARB2 | 1 | 1 | 1 |
| SCARF2 | 1 | 1 | 1 |
| SIGLEC1 | 1 | 1 | 1 |
| SKR3 | 1 | 1 | 1 |
| SMOC2 | 1 | 1 | 1 |
| SMPD1 | 1 | 1 | 1 |
| SPOCK1 | 1 | 1 | 1 |
| Siglec-9 | 1 | 1 | 1 |
| THY 1 | 1 | 1 | 1 |
| TMPRSS5 | 1 | 1 | 1 |
| TN-R | 0.94 | 1 | 1 |
| TNFRSF12A | 1 | 1 | 1 |
| TNFRSF21 | 1 | 1 | 1 |
| UNC5C | 0.83 | 1 | 1 |
| VWC2 | 0.62 | 1 | 1 |
| WFIKKN1 | 0.48 | 0.71 | 1 |
| gal-8 | 1 | 1 | 1 |
| sFRP-3 | 1 | 1 | 1 |

Table 3: Detectability values in nasal fluid, CSF and plasma using NULISAseq CNS panel. For each target, the proportion of samples with detection above LOD is given (1 = 100%).

|  | **NS** | **CSF** | **PLA** |
| --- | --- | --- | --- |
| ACHE | 1 | 1 | 1 |
| AGRN | 1 | 1 | 1 |
| ANXA5 | 1 | 1 | 1 |
| APOE | 1 | 1 | 1 |
| APOE4 | 0.51 | 0.5 | 0.44 |
| ARSA | 1 | 0.27 | 1 |
| Aβ38 | 0.94 | 1 | 1 |
| Aβ40 | 0.72 | 1 | 1 |
| Aβ42 | 0.15 | 1 | 1 |
| BACE1 | 0.21 | 1 | 1 |
| BASP1 | 0.4 | 0.14 | 0.81 |
| BD-MAPT | 1 | 1 | 0.38 |
| BD-pTau-181 | 1 | 1 | 1 |
| BD-pTau-217 | 1 | 1 | 0.62 |
| BD-pTau-231 | 1 | 1 | 0.75 |
| BDNF | 0.62 | 0.18 | 1 |
| CALB2 | 1 | 1 | 1 |
| CCL11 | 1 | 1 | 1 |
| CCL13 | 1 | 0.95 | 1 |
| CCL17 | 1 | 1 | 1 |
| CCL2 | 1 | 1 | 1 |
| CCL22 | 1 | 1 | 1 |
| CCL26 | 1 | 1 | 1 |
| CCL3 | 1 | 1 | 1 |
| CCL4 | 1 | 1 | 1 |
| CD40LG | 0.91 | 0.82 | 1 |
| CD63 | 1 | 1 | 1 |
| CHI3L1 | 1 | 1 | 1 |
| CHIT1 | 0.96 | 0.95 | 1 |
| CNTN2 | 0.81 | 1 | 1 |
| CRH | 0.34 | 1 | 0.94 |
| CRP | 1 | 1 | 1 |
| CSF2 | 1 | 1 | 1 |
| CST3 | 1 | 1 | 1 |
| CX3CL1 | 1 | 1 | 1 |
| CXCL1 | 1 | 1 | 1 |
| CXCL10 | 1 | 1 | 1 |
| CXCL8 | 1 | 1 | 1 |
| DDC | 1 | 1 | 1 |
| ENO2 | 1 | 1 | 1 |
| FABP3 | 1 | 1 | 1 |
| FCN2 | 0.77 | 0.5 | 1 |
| FGF2 | 1 | 0.27 | 1 |
| FLT1 | 1 | 1 | 1 |
| FOLR1 | 1 | 1 | 1 |
| GDF15 | 0.98 | 0.91 | 1 |
| GDI1 | 1 | 0.95 | 0.56 |
| GDNF | 0.43 | 0.27 | 0.94 |
| GFAP | 0.68 | 1 | 1 |
| GOT1 | 1 | 1 | 1 |
| HBA1 | 0.85 | 0.32 | 1 |
| HTT | 1 | 1 | 1 |
| ICAM1 | 1 | 1 | 1 |
| IFNG | 0.68 | 0.09 | 1 |
| IGF1R | 0.89 | 1 | 1 |
| IGFBP7 | 1 | 1 | 1 |
| IL10 | 1 | 1 | 1 |
| IL12p70 | 0.83 | 1 | 1 |
| IL13 | 0.83 | 0.32 | 0.94 |
| IL15 | 1 | 1 | 1 |
| IL16 | 1 | 1 | 1 |
| IL17A | 0.96 | 0.32 | 1 |
| IL18 | 1 | 1 | 1 |
| IL1B | 1 | 0.14 | 0.25 |
| IL2 | 0.77 | 0.77 | 1 |
| IL33 | 1 | 0.32 | 1 |
| IL4 | 0.53 | 0 | 0.88 |
| IL5 | 0.87 | 1 | 1 |
| IL6 | 1 | 1 | 1 |
| IL6R | 0.64 | 0.91 | 1 |
| IL7 | 1 | 1 | 1 |
| IL9 | 0.68 | 1 | 1 |
| KDR | 0.89 | 0.95 | 1 |
| KLK6 | 1 | 1 | 1 |
| MAPT | 1 | 1 | 1 |
| MDH1 | 1 | 1 | 1 |
| MME | 0.53 | 0.09 | 1 |
| MSLN | 1 | 1 | 1 |
| NEFH | 1 | 1 | 1 |
| NEFL | 0.96 | 1 | 1 |
| NGF | 1 | 1 | 1 |
| NPTX1 | 0 | 1 | 1 |
| NPTX2 | 0.94 | 1 | 1 |
| NPTXR | 0.85 | 1 | 1 |
| NPY | 1 | 1 | 1 |
| NRGN | 0.02 | 0.95 | 1 |
| Oligo-SNCA | 0.96 | 0 | 0.88 |
| PARK7 | 1 | 0.09 | 0.31 |
| PDGFRB | 1 | 1 | 1 |
| PDLIM5 | 0.6 | 0.41 | 0.75 |
| PGF | 1 | 1 | 1 |
| PGK1 | 1 | 0.55 | 1 |
| POSTN | 1 | 1 | 1 |
| PRDX6 | 1 | 1 | 1 |
| PSEN1 | 1 | 1 | 1 |
| pSNCA-129 | 1 | 0.14 | 1 |
| pTau-181 | 1 | 1 | 1 |
| pTau-217 | 1 | 1 | 0.94 |
| pTau-231 | 1 | 1 | 1 |
| pTDP43-409 | 1 | 0.23 | 0.69 |
| PTN | 0.89 | 1 | 0.06 |
| REST | 1 | 0.68 | 1 |
| RUVBL2 | 1 | 0.05 | 1 |
| S100A12 | 1 | 0 | 1 |
| S100B | 1 | 1 | 1 |
| SAA1 | 1 | 0.36 | 1 |
| SFRP1 | 1 | 1 | 1 |
| SFTPD | 1 | 0.95 | 1 |
| SLIT2 | 1 | 1 | 1 |
| SMOC1 | 1 | 1 | 1 |
| SNAP25 | 1 | 1 | 1 |
| SNCA | 1 | 1 | 1 |
| SNCB | 0.15 | 1 | 0 |
| SOD1 | 1 | 1 | 1 |
| SQSTM1 | 1 | 1 | 1 |
| TAFA5 | 1 | 1 | 1 |
| TARDBP | 1 | 0.95 | 1 |
| TEK | 1 | 1 | 1 |
| TIMP3 | 0.47 | 1 | 1 |
| TNF | 1 | 1 | 1 |
| TREM1 | 0.94 | 0.45 | 1 |
| TREM2 | 1 | 1 | 1 |
| UBB | 0.79 | 0 | 0.69 |
| UCHL1 | 1 | 0.86 | 0 |
| VCAM1 | 0.89 | 1 | 1 |
| VEGFA | 1 | 1 | 1 |
| VEGFD | 0.91 | 0.73 | 1 |
| VGF | 0.02 | 0.09 | 1 |
| VSNL1 | 1 | 1 | 1 |
| YWHAG | 1 | 0.23 | 0.44 |
| YWHAZ | 1 | 0.05 | 0 |
